# Brief interactive lifestyle preventive medicine video education in the primary care clinic: protocol for a randomized clinical trial

**DOI:** 10.1101/2025.03.20.25324341

**Authors:** Dylan Arroyo, Daniel Keyes, Ghadah W. Abdulshafi, Ali Jafri, Hemica Hasan, David Steinberger, Batoul Dabajeh, Alice Nassar, Anukul Karn, Rachel Nkrumah, Preanka Dhanoa, Kerby Shedden

## Abstract

**Introduction:** Preventive counseling improves long-term health, but primary care clinicians face time constraints limiting patient education. In a prior emergency department pilot study, a brief *passive* video trended towards increased patients’ willingness to change health behaviors. The current trial, conducted at the primary care clinic, evaluates the feasibility and impact of a short, *interactive* prevention video delivered during a primary care office visit.

**Methods and analysis:** We will conduct a prospective, randomized, non-blinded clinical trial among adult patients presenting to a primary care clinic. Immediately after their appointment, participants will be randomized to receive either the interactive prevention video with a practitioner or usual care. The video maintains engagement through simple in-video questions and adapts content based on patient responses (e.g., skipping smoking cessation for non-smokers). After the encounter, all participants will complete surveys assessing readiness and confidence to initiate lifestyle change including a transtheoretical “readiness ruler.” Survey items will be drawn from validated instruments, including the Gillespie & Lenz Readiness and Confidence to Participate in Lifestyle Activities Surveys, the Pittsburgh Sleep Quality Index (PSQI), and select questions from the U.S. Hospital Consumer Assessment of Healthcare Providers and Systems (HCAHPS) Survey. Patients will be asked for medical record access and permission for follow-up contact.

**Outcomes:** The primary outcome is index-visit Lifestyle Readiness and Confidence to Change scores. Secondary outcomes include satisfaction with the clinic visit, intention to change specific lifestyle behaviors, and healthcare use outside the clinic at 30 days and 6 months, as assessed via record review. A 3-12-month follow-up survey will assess self-reported lifestyle changes and new diagnoses.

**Statistical plan and sample size:** Analyses will follow intention-to-treat. Ordinal logistic regression will compare primary outcomes between arms, adjusting for Charlson Comorbidity Index, age, sex, gender, and prespecified predictors. Logistic regression will evaluate binary follow-up outcomes. Complete-case analyses will be used, and differential attrition will be reported. We plan to enroll approximately 350 patients.

**Trial registration:** https://ClinicalTrials.gov NCT06730737.

## Introduction

Preventive measures are critical in avoiding and limiting the severity of diseases. It is intuitive and widely accepted that preventing disease is much more potent than treating morbidities once they have developed. It has been estimated that health behaviors contribute as much as four-fifths of health outcomes, which can be attributed to behavioral and social factors, with clinical care only contributing a small proportion of influence [1,2]. This implies that the clinical encounter should always promote behavior changes, emphasizing prevention. However, contemporary healthcare is characterized by scheduling efficiencies emphasizing shorter times spent in direct contact between the provider and the patient. Direct observations of primary care practices have noted that prevention is under-emphasized [3]. Various reasons have been given for the lack of time for lifestyle change counseling. It is thus implicit that new methods are needed to educate patients about self-care and prevention that require less of a clinician’s time.

This project will evaluate the impact of a video-based educational intervention administered in the primary care clinic following the practitioner’s first encounter with the patient. It will also endeavor to examine the feasibility and efficiency of introducing patients and their accompanying family members to the importance of adopting changes in lifestyle behaviors.

Receptivity to health behavior changes has been extensively investigated. The transtheoretical model is a highly used measurement of receptivity to change and may be used to optimize intervention [4]. This model relates that there are six stages people fall into when adopting a more healthful lifestyle. The first stage is *pre-contemplation,* in which people have no plans to take action to change their behavior in the near future. This may be due to a need for more information or previous unsuccessful attempts to change. *Contemplation* occurs when people want to change their behavior within the next six months. They may be more aware of the benefits and costs, but this awareness can create trepidation and cause people to remain in this stage for extended periods. The third stage is *preparation*, defined as an individual’s readiness to change their behavior within the next month. They have a plan and are ready for an action-oriented program. The following stages include *action,* where changes are initiated within the last six months; *maintenance,* where people continue positive behaviors and work to prevent relapse; and *termination,* where people have no temptation to return to their old behavior. The transtheoretical model is often applied to preventive behavioral change. For example, it was reported that current smokers typically fall into a spread of 40% pre-contemplation, 40% contemplation, and 20% preparation [4]. This research suggests that when faced with the potential for serious adverse health outcomes, patients may be able to accelerate through some early stages, such as moving from pre-contemplation to contemplation or onto the preparation stage.

There have been previous attempts to implement prevention and education in the primary care clinic. These attempts include written after-visit summaries (AVS) shared with the patient. However, it has been reported that the AVS is not the most effective tool, considering that only 82.8% of patients recalled receiving an AVS, and 67.4% of patients consulted their AVS [5]. The information provided in the AVS is typically specific to the reason for that visit and includes contact information for a follow-up appointment. Another study measured whether patients given the AVS have better or worse glaucoma medication recall and revealed that 51.2% did not recall receiving any AVS. Those who recalled receiving one paradoxically had *lower* medication recall scores (remembering the name or color of the bottle or cap, treatment eye(s), and dosing regimen) than those who did not [6]. These results show that the intended goal of printed AVS, which is to enhance patient care, fails to be consistently achieved.

Moreover, there have been previous attempts at smoking cessation education in the primary care clinic. One example of these studies consisted of a training program focused on motivational interviewing to reduce smoking among adolescent patients. Follow-up assessments were conducted at 1, 3, and 6 months post-interview [7]. It was reported that abstinence rates at the 6-month follow-up were significantly higher in the motivational interview group compared to the standard advice control group [7].

Innovative approaches are needed to help patients absorb and retain the information required to properly ensure a healthy future. Showing a video on ways to enhance health while the patient is in the primary care provider (PCP) office could be an effective medium. In one study, patients who watched videos while they were waiting to be seen resulted in a higher proportion of patients rated their stay as “excellent” or “very good” (65%) in comparison to those who were not shown a video (58.1%) [8]. Those who received the video discharge instructions tested better on a post-test survey than the standard instructions group [9]. Another smaller study showed that after viewing an educational video on myocardial infarction, participants in the group shown the video had statistically significant improvement in the results of their knowledge assessment compared to the group not shown the video. The intervention group had a mean number of correct responses of 27 out of 37 questions as opposed to 20 in the control [10]. This supports the use of video as a practical approach to preventive education.

### Objectives

Primary objective: This study will evaluate the utility of presenting patients attending a primary care clinic with a focused video that will expose them to evidence-based education based on four of the core “pillars of preventive lifestyle health” promulgated by the American College of Lifestyle Medicine [11], including successful strategies for diet, exercise, sleep hygiene, and smoking cessation.

### Study hypotheses

We hypothesize that patients receiving the interactive video intervention will exhibit greater readiness and confidence to change lifestyle behaviors compared to patients in the control group, as measured post-intervention.

Null hypothesis (H₀): There is no difference in readiness and confidence to change lifestyle behaviors between the intervention group and the control group.

Alternative hypothesis (H₁): The intervention group will demonstrate significantly greater readiness and confidence to change lifestyle behaviors than the control group.

## Materials and methods

This will be a randomized, controlled, unblinded, two-armed superiority clinical trial. The study will be conducted according to the Standard Protocol Items: Recommendations for Interventional Trials (SPIRIT) Checklist. The study was approved by Trinity Health at Ann Arbor Institutional Review Board (IRB number: E-24-1117). The trial was prospectively registered in the https://Clinicaltrials.gov database (registration number: NCT06730737).

The study’s primary outcome will be the results from the Lifestyle Readiness to Change and Confidence to Change questionnaires, both of which use a *readiness ruler* [12]. The intervention will include four primary prevention domains promulgated by the American College of Lifestyle Medicine: diet, exercise, sleep hygiene, and smoking cessation.

Secondary aims include:

1. To evaluate the ability to implement a preventive education strategy in the primary care clinic setting
2. To assess the impact of age on willingness to change lifestyle behaviors
3. To determine those receiving the video concerning patient satisfaction, utilizing select Hospital Consumer Assessment of Healthcare Providers and Systems (HCAHPS) Survey questions, compared to the control group

### Eligibility

Inclusion criteria:

- All adults, including all ethnicities and genders, between the ages of 18 and 80 who come into the primary care clinic will be recruited to participate in the study.

Exclusion criteria:

- Non-English-speaking patient
- Unable or unwilling to consent to the study
- Unable or unwilling to hear a video on a smartphone or computer tablet
- The patient is in hospice care
- Patients with advanced dementia, incapable of answering the survey questions in the opinion of the person administering the survey

- Note: research associates will consult the patient’s primary practitioner or the physician members of the research team for any questions regarding dementia

Note:

- There are no relevant concomitant care and interventions that are permitted or prohibited during the trial.

### Participant enrollment and screening

Adult patients aged 18 to 80 will be recruited by a study team member or a research associate before their appointment or upon arrival at an Academic Internal Medicine ambulatory clinic. Please see the flow diagram for the randomization process and the table for the study process (Figure 1 and Table 1).

**Fig 1.**
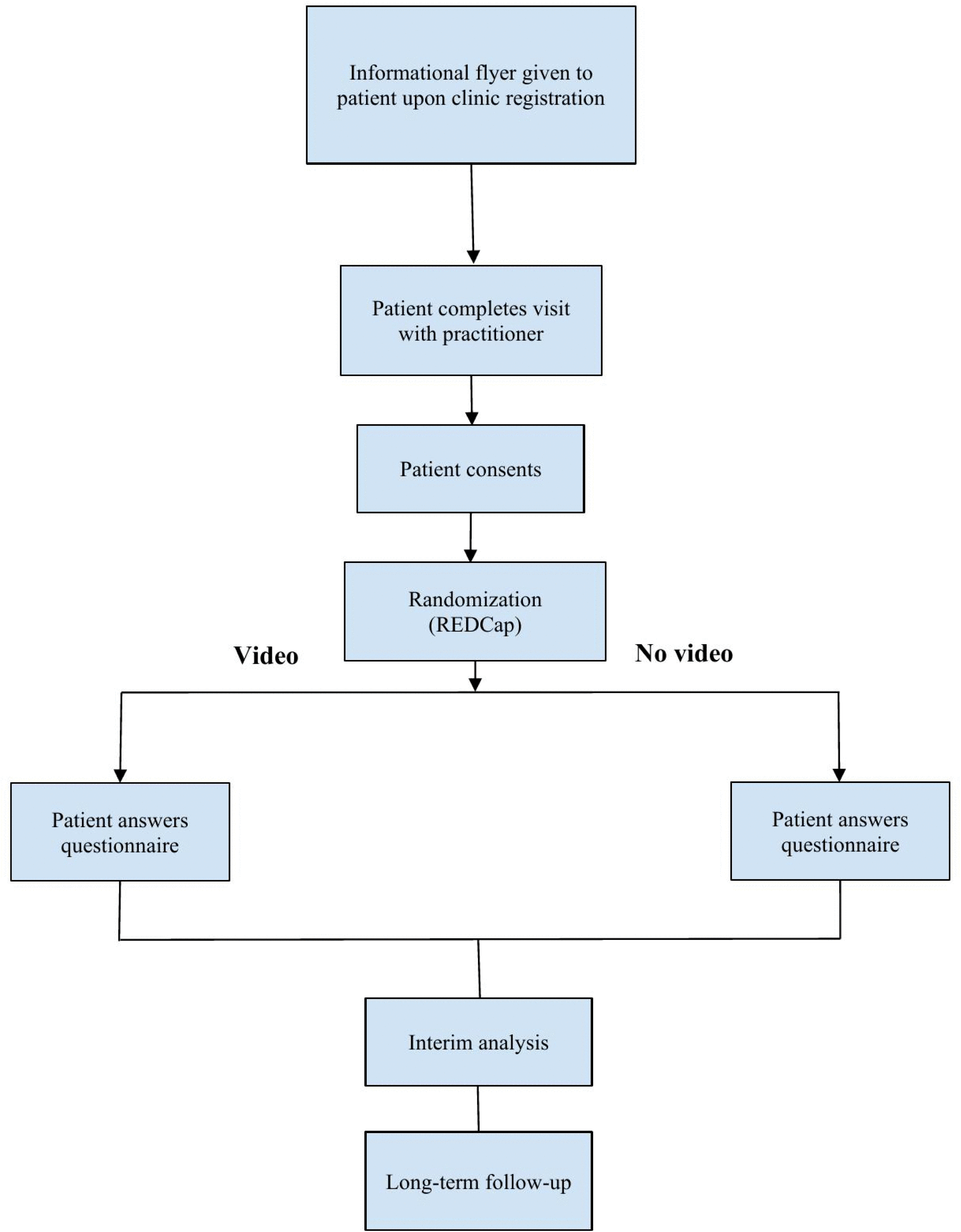
Flowchart for the Study Process. Note that all videos will use the interactive format. Patients will not be video recorded.

**Table 1.** Schedule of Enrollment, Interventions, and Assessments.

|  | STUDY PERIOD |  |  |  |  |
| --- | --- | --- | --- | --- | --- |
|  | Enrollment | Allocation | Post-allocation |  | Close-out |
| TIMEPOINT | -t<sub>1</sub> | 0 | t<sub>1</sub><br>(time 0) | t<sub>2</sub><br>(3-12<br>months) | t<sub>x</sub> |
| ENROLLMENT: |  |  |  |  |  |
| Eligibility Screen | X |  |  |  |  |
| Informed Consent | X |  |  |  |  |
| HIPAA Waiver | X |  |  |  |  |
| Randomization/Allocation |  | X |  |  |  |
| INTERVENTIONS: |  |  |  |  |  |
| Watch Video |  |  | X |  |  |
| Standard-of-care |  |  | X |  |  |
| ASSESSMENTS: |  |  |  |  |  |
| Baseline Assessment* | X |  |  |  |  |
| Primary Outcomes** |  |  | X |  | X |
| Secondary Outcomes*** |  |  | X |  | X |
| Follow-up Survey |  |  |  | X | X |

\*Sociodemographic, clinical, and lifestyle characteristics
\*\*Readiness/Confidence to Change questionnaires
\*\*\*Satisfaction with the primary care clinic visit and others

The randomization module within Research Electronic Data Capture (REDCap) will be used to allocate the video intervention. Those randomized for the intervention will be asked if they are interested and willing to watch a video about the clinic visit and their health. Those who agree to watch will then interact with a video featuring lifestyle changes relating to diet, exercise, sleeping, and smoking and why they are important for one’s overall health. The media is interactive because it contains questions for the user to answer at different points within the video sequence. The control group will not watch the video.

The study includes two surveys: the first will be administered in the clinic, and the second will be a follow-up survey administered upon phone contact with consenting patients. Target follow-up is planned for 3-12 months following the initial visit.

Whether receiving the video intervention or not, all participants will have provided verbal consent to answer the questionnaires. They will then fill out the questionnaire at the end of the video either alone, with the assistance of an accompanying relative, or with the assistance of a research team member to determine their stage of the transtheoretical model. A study identification number will be placed on the completed survey. The ID number and the patient’s identifying information will be noted in REDCap. Any patient who declines to participate will not have their patient health information (PHI) included in the study. The REDCap file containing the patients’ personally identifiable information will be kept on password-protected servers using the institution’s REDCap.

### Study procedures

Research associates and/or research team members will screen for eligible patients using a screening form based on the inclusion/exclusion criteria mentioned above. Results will be analyzed at two stages: first, with the results of the questionnaires administered as part of the initial visit, and second, after concluding the 3-12 month follow-up survey. The consent, educational video, and questionnaire will all be conducted while the patient is in the waiting room, after the clinician (physician or advanced practice provider) has completed the initial patient encounter, or after the provider contact has been completed.

The patients will fill out the composite questionnaire. This questionnaire is designed with the following criteria: prioritize using validated tools capable of self-administration. The Readiness-to-Change questionnaire evaluates patient readiness to implement ten lifestyle behaviors based on a 5-point rating scale. A rating of 5 represents having no interest in the suggested lifestyle change, which is consistent with the precontemplation stage of the transtheoretical model (TTM). A rating of 4 means an interest in implementing the lifestyle change in the next few months (contemplation stage of the TTM). A rating of 3 represents having the plan to implement the lifestyle change within the next month (preparation stage of the TTM). A rating of 2 represents having already started the lifestyle change within the past few months (action stage of TTM). A rating of 1 means having maintained lifestyle behavior changes for over 6 months (maintenance stage of TTM). The Confidence-to-Change questionnaire evaluates patient confidence in implementing the same lifestyle behaviors as the Readiness-to-Change questionnaire. However, the Confidence-to-Change questionnaire is based on a 3-point rating scale. Ratings are as follows: 3-very confident, 2-somewhat confident, and 1-not very confident [12]. The Confidence-to-Change questionnaire addresses any barriers or hesitancy that may impact patient confidence in implementing better lifestyle behaviors. Additional questions will include measures of satisfaction taken from the HCAHPS, specifically addressing their practitioner and visit.

Sometime after the visit (target: 3-12 months later), they will receive a phone follow-up call, asking them if they have adopted any lifestyle changes since watching the video, among other inquiries. We will use patients’ responses at this later stage (follow-up phase) to help determine how sustainable the preventive education intervention has been.

Randomized patients who participate in the study will receive the video intervention. In a previous pilot study, we investigated the impact of a simple instructional video on patients’ willingness to change in the emergency department setting [13]. The initial intervention was passive, requiring subjects to watch a video without interaction. The current study evaluates the practicality and impact of bringing a brief *interactive* educational video intervention to patients attending the primary care clinic visit.

All patients, including those randomized to “no video learning,” will be asked to complete a questionnaire. The questions will be administered orally or self-administered for those willing to participate using the tablet interface. Responses will be recorded using REDCap electronic data capture software and the related platform.

A sample size of approximately 350 was chosen based on feasibility and the expectation of sufficient power to detect moderate differences in readiness and confidence scores, guided by prior pilot study results. Due to the minimal risks in this study, a data monitoring committee is not considered necessary. An interim analysis will be conducted after 250 patients have been enrolled. It is anticipated that the total data collection will take six months. For those patients who agree to receive a call-back, contact will be attempted during the 3 to 12-month follow-up period. A research associate, a resident, or an attending physician will complete the follow-up by telephone or email correspondence with the subject. Up to 3 follow-up attempts will be made. Available follow-up data from the interactive electronic health record (EHR) will be obtained on all patients. The estimated time to view and interact with the video is approximately 10-15 minutes. The time required for responses to the survey component is estimated at 10-12 minutes.

The EHR will be used to determine several variables: age, race, ethnicity, sex and gender, BMI, and the Charlson Comorbidity Index (CCI). Michigan Data Analytics will assist in generating the patient’s CCI from ICD-10 codes. Information from the patient’s chart will only be obtained from those who provide informed consent.

The research team training will include informed consent, knowledge of the inclusion and exclusion criteria, using the EPIC EHR, and practice administering the study. The study will be performed using a convenience sample when team members are available to facilitate the study procedures.

Upon completion of the study, the results will be analyzed and presented at regional and national scientific meetings and for publication. The protocol will be submitted for publication prior to the initiation of the trial. Authorship of the resulting publication will follow the guidelines of the International Committee of Medical Journal Editors (ICMJE). No professional writers will be used for publication. No individual patient identifiers will be revealed during the abstract presentation or publication of the results. The data will be maintained in a password-protected electronic file on a hospital server using REDCap, a secure electronic data capture system.

## Statistical methods

The study team members will collect demographic data upon registration to the patient’s clinic. Patients from ages 18 to 80 will be included in the study. After informed consent, patients will be randomized to either the intervention or the control arm of the study. Only the interventional arm will interact with the video using a provided computer tablet which focuses on lifestyle preventive education. Control patients will not receive a video and will go directly to the follow-up questionnaire. Hence, both groups will receive the follow-up survey from research team members.

The Demographic data includes age, sex, gender, race, and insurance status. The patient’s BMI, Charlson Comorbidity Index, smoking status, responses to the social influences of health questionnaires, phone number, and frequency of ED visits will be documented from the patient’s EHR. Their zip code will also be collected for geographic and socioeconomic status analysis.

The study’s primary outcome is the readiness and confidence to change scores as a function of video exposure, measured on a continuous “readiness ruler.” [14].

Secondary outcomes will include readiness to change in specific domains, including exercise, smoking, diet, sleep hygiene, living an overall healthy lifestyle, satisfaction with the hospital, and satisfaction with the practitioner, dichotomized between those who watched the video and those who did not. Part of this data will be obtained directly from the patient’s medical record.

We will collaborate with Michigan Data Analytics, which will provide large-scale data abstraction to assist with obtaining the Charlson Comorbidity Index (CCI) for participating subjects. The remaining data will be collected using the Readiness-to-Change and Confidence-to-Change Lifestyle Questionnaires, the Pittsburgh Sleep Quality Index (PSQI), and U.S. Governmental HCAHPS Patient Satisfaction Survey.

Descriptive statistics will be used to summarize baseline characteristics by study arm, including means and standard deviations for continuous variables and proportions for categorical variables. All analyses will follow the intention-to-treat principle.

Outcomes collected at the clinic visit (e.g., readiness to change and confidence to change) will be compared between intervention and control groups using ordinal logistic regression, appropriate for ordinal outcome variables. As these measurements are taken after randomization and intervention (where applicable), no pre-intervention baseline measures will be available.

Follow-up outcomes, including receipt of additional care, adoption of lifestyle changes, and overall well-being, are recorded as binary or categorical variables (e.g., Yes/No/Don’t Know) and will be analyzed using logistic regression models. Clinic visit scores may be included as covariates to assess their association with follow-up outcomes.

A complete-case approach will be used for all primary and secondary outcome analyses. Differences in loss to follow-up between study arms will be reported, and characteristics of completers and non-completers will be compared to assess potential attrition bias. Multiple imputation is not planned due to the structure of data collection involving a single follow-up time point.

Separate regression models will be used to analyze outcomes collected at the clinic visit and follow-up. Given the design of the study, which includes a single post-intervention follow-up and no pre-intervention baseline, repeated measures modeling will not be used in the primary analysis. Secondary analyses may include generalized estimating equations or other repeated measures approaches to assess the consistency of findings.

### Data confidentiality and storage

All team members will be trained in HIPAA and safe data handling practices. To maintain patient confidentiality and privacy, the data will be maintained on a password-protected secure server and using the secure, password-protected REDCap platform.

### Data and safety monitoring plan

The principal investigator (PI) is responsible for this project’s conduct and shall ensure that all work and services described, or incidental to those described, are conducted following accepted standards of medical practice. The PI and co-investigators will maintain all master subject lists, and all data collected will be maintained in a password-protected secure server and reviewed at regular intervals to ensure adherence to the investigational procedures outlined in the protocol.

The approved research team members will obtain information regarding the patient’s lifestyle and demographic habits. Data will be accepted from primary care clinic records and charts. Each data abstractor will use a pre-formulated data collection tool to enhance the uniformity of information. Statistical analysis for inter-observer variability will be conducted on a subset of duplicate charts to ensure the quality of the data abstracted and used for the study.

Meetings will be held at regular intervals to discuss data management, general IRB compliance, and other study aspects and to analyze the collected data. The PI will evaluate data quality and ensure that data evaluation is performed frequently. All data will be documented and retained in the study files.

## Regulatory requirements

### Informed consent

All patients will be consented. Informed consent will be obtained before the randomization of subjects. If they approve, the research team member will administer the informed consent form prior to randomization.

Those who approve to watch the interactive video will be given a QR code to watch on their smartphone or a tablet with a link to the video, which the patient or the research team member may start.

### Participant privacy

Subject confidentiality will be maintained using a randomly generated participant identification number/code. The participant identification number will be stored with the patient’s identifying information, such as name and room number, for verification purposes. Identifying information will be kept in a password-protected, secure REDCap file on a password-protected server. Such information will only be accessible to the research team trained in research ethics (CITI training) and HIPAA compliance.

Using personally identifiable data is necessary to determine eligibility, abstract chart data, and follow up with patients. All patient information collected will be stored in a password-protected secure server using REDCap and only accessed when necessary by authorized research associates.

### HIPAA

All parties involved shall maintain the confidentiality of PHI to ensure compliance with privacy regulations, including the Health Insurance Portability and Accountability Act of 1996 (HIPAA). The study team will consist of only personnel trained in research ethics (CITI training) and HIPAA compliance.

Patient eligibility will be determined by screening in Epic EHR. We will request a waiver of HIPAA authorization for this portion of the study and access to the patient’s medical record.

### Unanticipated problems and adverse events

This project is considered to be minimal risk, such as loss of protected health information and discomfort of answering questions. Any unanticipated problems will be promptly reported to the PI, project’s research coordinator, and the IRB. If necessary, the project will be discontinued until resolution in consultation with Trinity Health, Ann Arbor IRB. This trial involves preventive health education and does not involve any interventions that carry a risk of physical harm. Therefore, provisions for ancillary and post-trial care related to trial participation, as well as compensation for harm, are not applicable.

## Limitations and assumptions

Since this is planned as a single-institution study, this will limit generalizability.

## Data availability

This manuscript describes a study protocol. No data has been generated or analyzed at this stage. Upon completion of the study, all de-identified individual-level data underlying the findings will be made publicly available in a recognized public repository in accordance with Trinity Health IRB policies and participant consent provisions.

Study materials such as the survey instruments (including the Readiness-to-Change and Confidence-to-Change questionnaires, Pittsburgh Sleep Quality Index [PSQI], and HCAHPS items) and the video are available from the corresponding author upon reasonable request and will be made publicly available upon peer-reviewed publication.

